# Evaluation of the disease outcome in Covid-19 infected patients by disease symptoms: a retrospective cross-sectional study in Ilam Province, Iran

**DOI:** 10.1101/2020.11.10.20228908

**Authors:** Jamil Sadeghifar, Habib Jalilian, Khalil Momeni, Hamed Delam, Tadesse Sheleme, Ayoub Rashidi, Fariba Hemmati, Shahab Falahi, Morteza Arab-Zozani

## Abstract

**Background:** novel coronavirus disease-19 (COVID-19) announced as a global pandemic in the year 2020. With the spread of the disease, a better understanding of patient outcomes associated with their symptoms in diverse geographic levels is vital. We aimed to analysis clinical outcomes of COVID-19 patients by disease symptoms in Ilam province of Iran.

**Methods:** This is a retrospective study. Data were collected from integrated health system records for all hospitals affiliated to Ilam University of Medical Sciences between 26 Jan 2020 and 02 May 2020. All patients with definite positive test were enrolled in this study. We used descriptive analyses, chi-square test and binary logistic regression to analyze the data using SPSS version 22.

**Results:** The mean age was 46.47±18.24 years. Of 3608 patients, 3477 (96.1%) were discharged and 129 (3.9%) were died. 54.2% of the patients were male and were in the age group of 30-40 years old age. Cough, sore throat, shortness of breath or difficulty breathing and fever or chills were the most common symptoms. People with symptoms of shortness of breath, abnormal radiographic findings of the chest, and chest pain and pressure were relatively more likely to die. Based on the findings of binary logistic regression probability of death in people who showed shortness of breath, abnormal chest radiographic findings and chest pain was 1.34, 1.24 and 1.32 times higher than those who did not show these symptoms, respectively.

**Conclusion:** Our study provides evidence that presentation of some symptoms does significantly impact on outcomes of patients infected with SARS-CoV-2. Early detection of symptoms and proper management of outcomes can reduce mortality in patients with COVID-19.

## Background

The first cases of an unknown pneumonia were reported in late December 2019 from Wuhan, China [1]. After a while, a novel Coronavirus, severe acute respiratory syndrome coronavirus 2 (SARS-COV-2), was identified as the causative agent and was subsequently named novel Coronavirus disease 2019 (COVID-19) by the World Health Organization (WHO) [2]. A few months after the onset of the disease, the WHO on 11 March 2020 has declared the disease outbreak a global pandemic [3]. More than 49 million cases worldwide and 600,000 cases in Iran have been reported so far, of which more than 1 million deaths have been recorded in the world, which is a significant statistic compared to previous pandemics [4, 5]. To date (11.07.2020), according to statistics, Iran ranks as 14^th^ in terms of the number of cases and the 9^th^ country in terms of the number of deaths due to COVID-19 [5]. Considering the specific features of this disease and its increasing trend in the world and especially in Iran, identifying the epidemiological characteristics of patients and their clinical features is very important in making appropriate decisions that can ultimately help control the epidemic [6]. Main features of COVID-19 have been previously reported [7, 8], but it is also important to look at the characteristics of individuals in certain populations. The epidemiological characteristics of patients in a particular region can be different from other parts of the world, so knowledge of these epidemiological and clinical characteristics can be useful to local authorities to provide the necessary facilities and to decide on disease control [9-11]. Then, this study aimed to evaluate the disease outcome in COVID-19 infected patients by disease symptoms in Ilam province, west of Iran.

## Methods

### Study design, patients and data collection

The present study is a retrospective descriptive cross-sectional study. Data were collected at the all hospitals that affiliated to Ilam University of Medical Sciences. All suspected individuals who have been referred to and tested for COVID-19 and his/her information registered in integrated health system records of Iran called SIB and related to Ilam provinces, between 26 Jan 2020 and 02 May 2020 with definite positive test were enrolled in this study. The proposal was approved by the ethics committee of Ilam University of Medical Sciences (No. IR.MEDILAM.REC.1399.043). The extracted information were as follows: gender, age, national code, date of admission and hospitalization, sign and symptoms, contact history, type of comorbidities, test results, and final outcome. Positive cases of COVID-19 were confirmed by SARS-CoV-2 real-time reverse transcriptase–polymerase chain reaction (RT-PCR). Data were collected at the 4 hospitals that provided care for these patients. All extracted information is kept confidential.

### Statistical analysis

Retrieved data were recorded into MicrosoftExcel (version 13) and analyzed. The SPSS version 22.0 (SPSS 22.0; SPSSInc, Chicago, IL, USA) were used. Two researchers independently reviewed the collected data for accuracy. Descriptive analyses of the variables were conducted as mean (Mean±SD) or number (%). Chi-square test was used to evaluate the effect of demographic variables (age, sex, etc.) and underlying diseases / symptoms on the outcome of the infected patients. Also, the cumulative effect of all independent variables on disease outcome was investigated using binary logistic regression.

## Results

The mean (±SD) of age the participants was 46.47±18.24. Out of 3608 patients, 3477 (96.1%) were discharged and 129 (3.9%) died. Most of the patients were male and were in the age group of 30-40 years old age Table 1. Cough, sore throat,shortness of breath or difficulty breathing and fever or chillswere the most common symptoms, respectively Table 2.

**Table 1:**
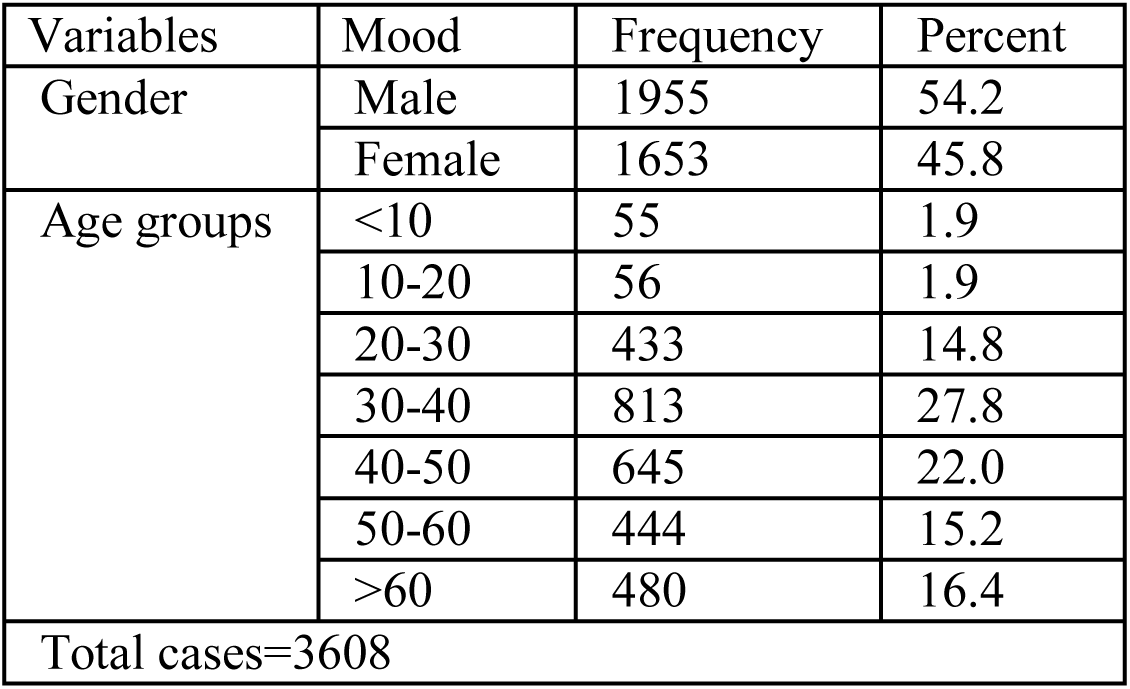
frequency of gender and age groups

| Variables | Mood | Frequency | Percent |
| --- | --- | --- | --- |
| Gender | Male | 1955 | 54.2 |
|  | Female | 1653 | 45.8 |
| Age groups | <10 | 55 | 1.9 |
|  | 10-20 | 56 | 1.9 |
|  | 20-30 | 433 | 14.8 |
|  | 30-40 | 813 | 27.8 |
|  | 40-50 | 645 | 22.0 |
|  | 50-60 | 444 | 15.2 |
|  | >60 | 480 | 16.4 |
| Total cases=3608 |  |  |  |

**Table 2:** frequency of disease symptoms

| Symptoms | Frequency | Percent |
| --- | --- | --- |
| Cough | 1465 | 40.6 |
| Shortness of breath or difficulty breathing | 949 | 26.3 |
| Sore throat | 931 | 25.8 |
| Fever or chills | 866 | 24.0 |
| Muscle or body aches | 401 | 11.1 |
| Headache | 339 | 9.4 |
| General weakness | 237 | 6.6 |
| Nausea or vomiting | 120 | 3.3 |
| Diarrhea | 115 | 3.2 |
| Abnormal findings of chest radiography | 98 | 2.7 |
| Conjunctivitis | 91 | 2.5 |
| New confusion | 85 | 2.4 |
| Persistent pain or pressure in the chest | 53 | 1.5 |
| Stomach ache | 52 | 1.4 |
| Congestion or runny nose | 22 | .6 |
| External pharynx | 18 | .5 |
| Eye redness | 9 | .2 |
| Total cases=3608 |  |  |

According to the results of Chi-square and Fisher‘s Exact Test, people with symptoms of shortness of breath, abnormal chest radiograph findings, and chest pain and pressure are relatively more likely to die. On the other hand, people with symptoms of sore throat, headache, diarrhea, contusions, and muscle pain were relatively less likely to die (Table 3).

**Table 3:** association between disease symptoms and disease outcome

| Symptoms | Discharge | Death | X<sup>2</sup> | P value |
| --- | --- | --- | --- | --- |
| Shortness of breath or difficulty breathing | 22.8% | 56.6% | 77.26 | <.0001 |
| Abnormal findings of chest radiography | 1.7% | 10.1% | 42.81 | <.0001 |
| Persistent pain or pressure in the chest | 1.4% | 4.7% | 8.52 | .004 |
| Sore throat | 27.0% | 11.6% | 15.02 | <.0001 |
| Headache<sup>*</sup> | 10.1% | 2.3% | 8.45 | .001 |
| Diarrhea<sup>*</sup> | 3.5% | 0.0% | 4.66 | .02 |
| Muscle or body aches | 12.0% | 3.9% | 7.89 | .005 |
| Fever or chills | 24.7% | 17.8% | 3.17 | .075 |
| Eye redness<sup>*</sup> | 0.3% | 0.0% | .36 | 1.00 |
| Cough | 39.3% | 39.5% | .002 | .96 |
| General weakness | 6.2% | 9.3% | 2.004 | .15 |
| Nausea or vomiting<sup>*</sup> | 3.5% | 2.3% | .48 | .62 |
| Conjunctivitis<sup>*</sup> | 2.7% | 2.3% | .059 | 1.00 |
| New confusion<sup>*</sup> | 2.6% | 0.8% | 1.65 | .37 |
| <sup>*</sup>Abnormal lung sound | 0.1% | 0.8% | 3.45 | .18 |
| Stomach ache<sup>*</sup> | 1.6% | 0.0% | 2.105 | .26 |
| Congestion or runny nose<sup>*</sup> | 0.6% | 0.0% | .81 | 1.00 |
| External pharynx<sup>*</sup> | 0.6% | 0.0% | .73 | 1.00 |

<sup>\*</sup>In cases where the number of observations was less than 5, Fisher's exact test was used instead of Chi-square test

The results of the binary logistic regression model also confirmed the above findings. Accordingly, shortness of breath, abnormal chest radiograph findings, and chest pressure and pain predict the outcome of death in patients. Patients who showed symptoms of shortness of breath, abnormal findings on chest radiography, and chest pain and pressure were more likely to die than those who did not show these symptoms (1.34, 1.24, and 1.32, respectively) (Table 4).

**Table 4:** binary logistic regression of disease outcome by disease symptoms

| Predictors | B | P value | Wald | Exp(B) |
| --- | --- | --- | --- | --- |
| Shortness of breath or difficulty breathing | 1.344 | <.0001 | 49.940 | 3.834 |
| Persistent pain or pressure in the chest | 1.242 | .011 | 6.523 | 3.464 |
| Abnormal findings of chest radiography | 1.320 | <.0001 | 13.541 | 3.745 |
| Sore throat | -.675 | .02 | 5.452 | .509 |
| Headache | -1.177 | .057 | 3.609 | .308 |
| Body pain | -.880 | .070 | 3.289 | .415 |
| Fever and chill | -.302 | .212 | 1.555 | .739 |
| Cough | -.192 | .334 | .931 | .825 |
| General weakness | .457 | .170 | 1.880 | 1.580 |
| New confusion | -1.056 | .305 | 1.051 | .348 |
| Congestion or runny nose | -17.239 | .998 | .000 | .000 |
| Diarrhea | -17.786 | .996 | .000 | .000 |
| Nausea and vomiting | .404 | .511 | .432 | 1.498 |
| Stomach ache | -17.092 | .997 | .000 | .000 |
| Conjunctivitis | .932 | .143 | 2.146 | 2.539 |
| External pharynx | -16.458 | .999 | .000 | .000 |
| Redness eyes | -1.186 | 1.000 | .000 | .305 |
| Constant | -3.435 | .000 | 437.435 | .032 |

## Discussion

In the present study, which was designed to evaluate the outcome of the disease in patients with COVID-19 according to the symptoms of the disease, it was found that from January to May 2020 in Ilam province in western Iran, 3608 definitive cases of COVID-19 have been diagnosed. Of these, 3.9% had died of the disease. Studies around the world show a significant increase in COVID-19 cases and deaths worldwide [12]. A study conducted in southern Iran showed that COVID-19 mortality was about 8% [13], and a meta-analysis study showed that overall COVID-19 mortality was about 5%. [14], which shows a higher percentage than the results of the present study. It seems that the differences in the demographic characteristics of the patients as well as the severity of the disease of the patients participating in the studies have led to the reporting of different percentages of disease mortality.

In the present study, the majority of infected cases were men.Women appear to be less susceptible to viral infections because of their protection against the X chromosome and sex hormones that play a key role in their innate immunity [15]. Also, the age group of 30-40 years had the highest frequency of infected cases. A similar study in Iran showed that the age group of 50-60 years has the highest incidence of COVID-19, while the highest fatality rate of the disease with 19.27 and 14.85% is related to age groups with more than 70 years [6]. It seems that with increasing age, the probability of death due to COVID-19 also increases. As in the study of Zhou et al., It was found that patients who died due to COVID-19 had a significantly higher mean age (about 69 years) than those people who survived [16].

The results of logistic regression model in the present study showed that symptoms such as shortness of breath, pain and pressure of chest, abnormal findings of chest radiography and sore throat are the most important factors in predicting the outcome of death in patients with COVID-19.According to the results of this study, people who had a higher percentage of shortness of breath, chest pain and pressure, and abnormal chest radiographic findings were more likely to die than other patients. However, in terms of other symptoms of the disease, no significant difference was observed between the two groups (dead or survived).The study by Chang et al. showed that symptoms such as fever and chills significantly contribute to the progression of the disease to its severe stage [17]. Another study by Lian et al. showed that patients over 60 years of age and shortness of breath were significantly more likely to develop the more severe clinical form of COVID-19 than other patients [18]. Another similar study found that 100% of patients with COVID-19 whose lungs were damaged were died [19].The study by Li et al. showed that the prevalence of symptoms such as cough, sputum, chest pain, and shortness of breath in patients with severe or acute COVID-19 was significantly higher than in normal patients. This study found that patients with severe disease had a CT scan score higher than 7 compared to patients with normal condition [20]. On the other hand, a review study by GalloMarin et al. showed that hypoxia and specific CT scan findings indicate extensive lung involvement, with increased disease severity or death [14].Chest imaging results in one study showed that the presence of pulmonary fibrosis and older age was associated with increased patient transfer to the intensive care unit (ICU) [21]. However, in the study of Guan et al., It was found that 20.1% of all patients with COVID-19 who had a positive RT-PCR test and also had symptoms of the disease were normal in terms of chest CT scan [7]. It is therefore important to note that CT scan alone cannot be a diagnostic criterion for COVID-19 [22].

Another study by Chen et al. Showed that fever and shortness of breath were significantly higher in patients who died of COVID-19 than in the survived group [23]. Another similar study found that the prevalence of symptoms such as shortness of breath, chest tightness, fatigue, muscle pain and dizziness was higher in patients with severe or critical COVID-19.Also, only in terms of shortness of breath, there was a significant difference between the group that died and the group that survived. The percentage of shortness of breath in the group that died of COVID-19 was significantly higher than the other group [24].

## Conclusion

The results of the present study showed that symptoms such as shortness of breath, pain and pressure of chest, abnormal findings of chest radiography and sore throat are the most predictors of the outcome of death in patients with COVID-19. Patients who had a higher percentage of shortness of breath, chest pain and pressure, and abnormal chest radiographic findings were more likely to die than other patients.

## Data Availability

Data is available in case of request.

## List of abbreviations

SARS-COV-2: severe acute respiratory syndrome coronavirus 2
COVID-19: novel Coronavirus disease 2019
RT-PCR: real-time reverse transcriptase–polymerase chain reaction
WHO: World Health Organization
ICU: intensive care unit

## Ethics approval and consent to participate

This study was approved by the ethics committee of Ilam University of Medical Sciences (No. IR.MEDILAM.REC.1399.043). All data extracted from Health information system and the authors are committed to maintaining the confidentiality of the data.

## Consent for publication

Not applicable

## Availability of data and materials

The datasets analyzed during the current study are available in the integrated health system records of Iran called SIB and the Ilam University of Medical Sciences.

## Authors’ contributions

MAZ, KhM, JS designed the study. SF, AR and FH collected the data. HJ and MAZ analyzed and interpreted the data. MAZ and HD drafted the manuscript. TSh revised the manuscript. MAZ and JS supervised the manuscript writing, reviewed the manuscript for intellectual content and scientific integrity. All the authors reviewed and approved the manuscript before publication.

## Acknowledgements

Not applicable

